# Miniaturized Four-Dimensional Functional Ultrasound for Mapping Human Brain Activity

**DOI:** 10.1101/2025.08.19.25332261

**Authors:** Luuk Verhoef, Sadaf Soloukey, Geert Springeling, Adriaan J. Flikweert, Boris Lippe, Arend J. de Jong, Nikola Radeljic-Jakic, Michiel Baas, Jason Voorneveld, Arnaud J.P.E. Vincent, Pieter Kruizinga

**Affiliations:** Brain Echo Lab, Department of Neuroscience, Erasmus MC, 3015 GD, Rotterdam, Netherlands; Department of Neurosurgery, Erasmus MC, 3015 GD, Rotterdam, Netherlands; Department of Experimental Medical Instrumentation, Erasmus MC, 3015 GD, Rotterdam, Netherlands; Oldelft Ultrasound, 2629 JG, Delft, Netherlands; Electronic Instrumentation Laboratory, Delft University of Technology, 2628 CD Delft, Netherlands; Department of Microelectronics, Delft University of Technology, 2628 CD, Delft, Netherlands

## Abstract

Real-time brain monitoring for neurosurgery and neuroscience research of natural behaviors demands portable imaging with high spatiotemporal resolution. Current technologies cannot simultaneously achieve the resolution, mobility, and real-time performance required. Here we present a miniaturized four-dimensional functional ultrasound system capturing volumetric brain hemodynamics in real-time using 3072 transceivers controlled by custom application-specific integrated circuits. The device achieves 450 Hz volumetric imaging up to 8 cm depth while maintaining a form factor suitable for direct cortical placement and potential sub-cranial implantation. We validated this technology across three clinical scenarios: through skull prosthesis, cranial defect, and during neurosurgery. The system mapped somatotopic finger representations and achieved decoding of individual finger movements during piano playing using machine learning, demonstrating single-trial detection. This portable platform establishes a new approach for brain monitoring bridging laboratory neuroscience and clinical applications, enabling research in natural behavioral settings, providing surgeons real-time hemodynamic feedback, and advancing brain-computer interface development.

---

The human brain operates through intricate patterns of activity that unfold across space and time, yet our ability to observe these dynamics in real-world settings remains severely constrained. Neurosurgeons operating near eloquent brain areas require real-time feedback about functional tissue to minimize post-surgical deficits, but no current technology provides this capability during surgical procedures. Similarly, understanding how the brain functions during natural behaviors, from playing musical instruments to social interactions, demands portable imaging that can capture neural activity outside traditional laboratory constraints. These critical applications reveal the need for real-time, high-resolution brain monitoring that operates in environments where current neuroimaging methods cannot.

Functional magnetic resonance imaging (fMRI) provides excellent spatial coverage but suffers from low spatial resolution (∼1 mm), slow temporal sampling (∼2 Hz), and confines subjects within restrictive scanner environments that require massive infrastructure (*1, 2*). Portable techniques like electroencephalography (EEG) and electrocorticography (ECoG) lack the spatial precision or depth resolution needed for detailed cortical mapping during complex behaviors or surgical guidance (*3–6*). These constraints prevent applications for both real-time surgical decision-making and the study of brain function during ecologically valid behaviors.

Functional ultrasound imaging (fUSi) offers a promising alternative by detecting hemodynamic changes associated with neural activity at sub-millimeter spatial resolution and sub-second temporal precision (*7, 8*). Previous work has demonstrated fUSi’s potential across multiple clinical contexts: intra-operative mapping during neurosurgical procedures (*9–11*), real-world brain monitoring through sonolucent skull prostheses during natural activities like walking and guitar playing (*12, 13*), neonatal brain connectivity monitoring through the fontanel (*14, 15*), and even brain-computer interface applications in non-human primates (*16,17*). While four-dimensional fUSi has been demonstrated in animal models (*18*), human applications have been fundamentally limited by two-dimensional imaging approaches that can only visualize a single 2D plane of the brain. As illustrated in Figure 1A, conventional 2D fUSi imaging inherently cannot capture the volumetric nature of cerebral dynamics, such as seen in the three-dimensional organization of the sensorimotor cortex. This limitation becomes particularly critical for surgical applications where out-of-plane vascular or functional activity affects decision-making, and for brain-computer interfaces where comprehensive spatial coverage determines decoding fidelity. The step towards 4D fUSi in humans was held back primarily by the lack of suitable ultrasound transducer technology and processing software to deal with the challenging data rates for real-time imaging feedback.

**Figure 1:**
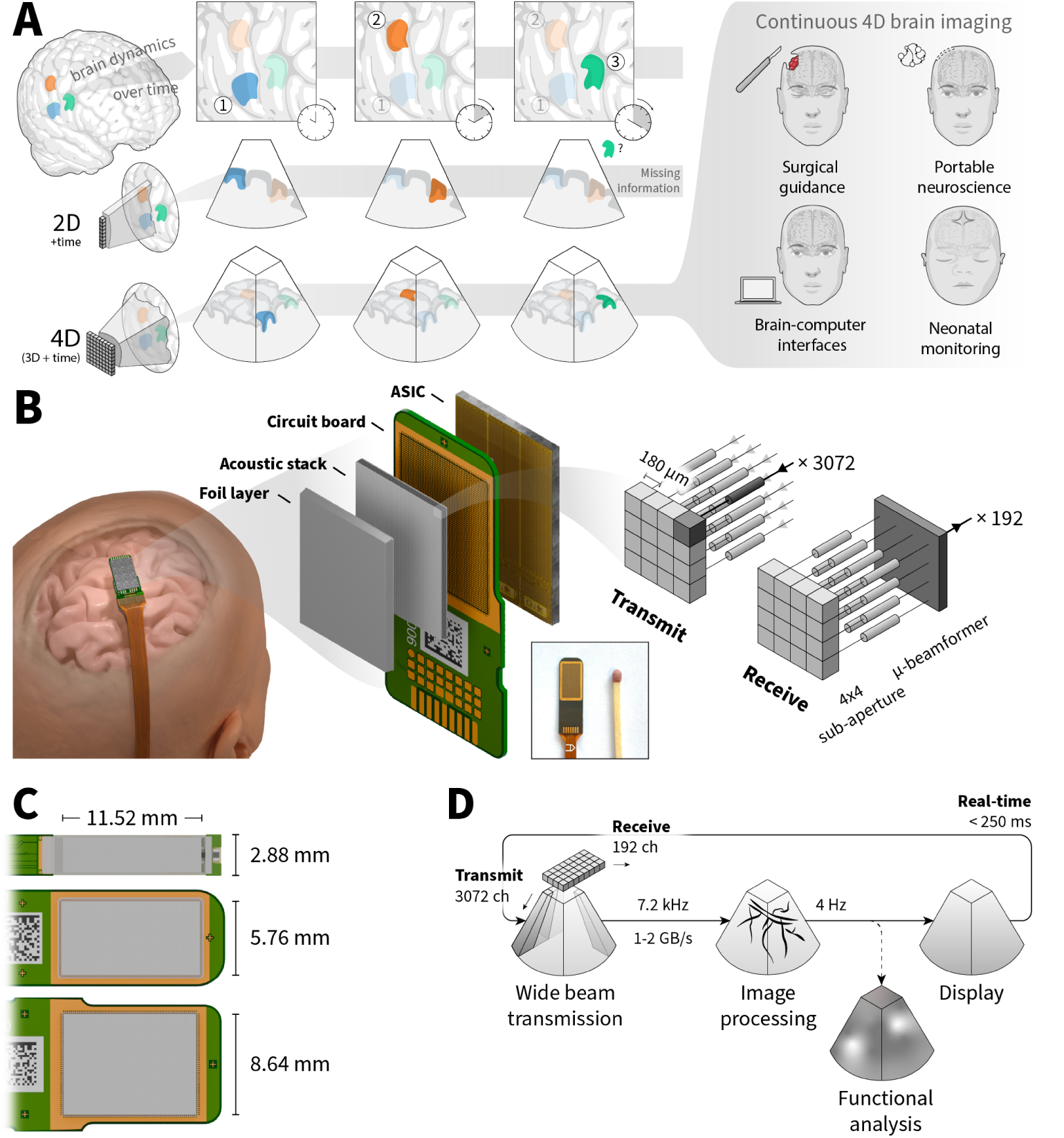
Miniaturized four-dimensional functional ultrasound overcomes 2D imaging limitations. (**A**) Conventional 2D fUSi misses volumetric activation patterns (green region outside imaging plane), while a 4D probe captures complete cortical dynamics, enabling four key applications: intraoperative monitoring, mobile brain imaging for neuroscience experiments, neonatal brain monitoring through the fontanel, and fUSi-based brain-computer interfaces. (**B**) System overview showing the probe on a subject’s head (left), exploded view of the miniaturized probe architecture with 3072 piezoelectric elements and three tiled application-specific integrated circuits (ASICs, middle), and micro-beamforming architecture within each 4×4 element group achieving 16:1 channel reduction (right). (**C**)Probe configurations using one, two, and three tiled ASICs with aperture dimensions. (**D**) Real-time processing pipeline achieving < 250 ms latency from acquisition to hemodynamic visualization.

Here, we present the first four-dimensional (4D) functional ultrasound imaging system for portable and real-time human brain monitoring. Our approach overcomes previous limitations through three key advances: a miniaturized 3072-element ultrasound probe with custom application-specific integrated circuits (ASICs), high frame rate acquisition sequences optimized for hemodynamic sensitivity, and GPU-accelerated reconstruction algorithms. The complete system captures brain activity up to 5-8 cm depth across a 60^◦^ × 60^◦^ field of view (FOV) at 1-10 Hz temporal resolution, while maintaining an 11.52 × 8.64 mm footprint suitable for integration into surgical workflows and future intracranial implantation. This miniaturization addresses multiple clinical and scientific needs simultaneously: enabling access through the small craniotomies typical of modern neurosurgery, through natural acoustic windows like the neonatal fontanel, and providing the mobility essential for studying brain function during unconstrained behaviors.

We first describe the probe design and ASIC architecture enabling volumetric imaging from a miniaturized platform, then demonstrate hemodynamic imaging performance across three human subjects with acoustic brain access: during neurosurgery, through a cranial defect, and via a sonolucent skull prosthesis. Our functional validation ranges from mapping known somatotopic organization to achieving single-trial decoding of individual finger movements of a subject playing the piano. These results establish 4D fUSi as a practical tool for applications where current imaging modalities fall short: providing surgeons with real-time hemodynamic feedback during procedures and enabling neuroscience research in natural behavioral settings.

## Programmable miniature 4D ultrasound probe

Developing a miniature probe for high frame rate (ultrafast) volumetric brain imaging required overcoming fundamental engineering constraints. Conventional probes for two-dimensional imaging typically employ a linear array of 64-192 densely packed elements, with configurations spaced at less than half the acoustic wavelength (phased arrays) providing the greatest flexibility in beam steering. On the other hand, volumetric imaging requires a matrix array where element count scales quadratically, quickly necessitating thousands of elements for any practical application. This dramatic increase in channel count far exceeds the sampling capabilities of standard ultrasound systems, which typically support only 128-256 simultaneous channels.

To bridge this gap, we developed a custom application-specific integrated circuit (ASIC) positioned directly behind the acoustic elements, performing initial signal processing with 16:1 channel reduction (Figure 1A). The ASIC architecture supports a pitch-matched connection of 64 × 16 elements spaced at 180 × 180 𝜇m (a phased array), with integrated transmit and receive circuitry for each element. Our probe contains three tiled ASICs, yielding 3072 addressable elements operating at 5 MHz center frequency with >50% bandwidth, compressed into an 11.52 × 8.64 mm footprint (Figure 1B). The transmit capabilities include programmable waveform generation and beam steering, while the receive pathway incorporates low-noise signal amplification, time-gain-compensation and delay-and-sum micro-beamforming. Groups of 4 × 4 elements are combined into single output channels using individually programmable delays, reducing the channel count from 3072 to 192, which is easily manageable with modern high-end ultrasound systems.

Despite its compact footprint, the probe can achieve a substantial imaging field of view by transmitting diverging beams. With elements spaced at less than half the acoustic wavelength and the ASIC enabling precise, and independent delay control for each transmit element, spherically diverging wavefronts can be generated to insonify an angular FOV much larger than the probe’s aperture.

While ASICs have been employed previously to reduce channel count in ultrasound probes (*19, 20*), designs have typically imposed constraints on imaging sequences that prevented the high frame rates required for functional ultrasound applications. Our ASIC architecture uniquely enables the kilohertz pulsing rates (approaching 10 kHz) required for volumetric ultrafast imaging sequences, achieved through efficient control interfaces and optimized micro-beamforming that maintains signal fidelity at these rates.

## High resolution volumetric imaging of brain hemodynamics

Detecting brain activity through ultrasound demands extraordinary sensitivity to capture the hemodynamic variations related to neural activation. Neurovascular coupling, the phenomenon where increased neural activity drives local blood flow changes, forms the physiological basis for both fMRI and fUSi (although fUSi measures blood volume rather than blood oxygenation) (*2, 7, 21, 22*). The hemodynamic response of interest occurs primarily in the microvasculature, with baseline flow velocities of ∼ 1 mm/s (*7, 23*). The fundamental challenge for a fUSi system is therefore to attain sufficient sensitivity to distinguish this slow-flowing blood from a much stronger background signal from the surrounding slow-moving (brain) tissue.

Ultrafast Doppler imaging achieves this sensitivity by transmitting unfocused beams and performing all focusing computationally, enabling frame rates approaching 10 kHz, over 10-fold higher than traditional imaging (*8*). This temporal oversampling enables statistical separation of flow from moving tissue based on their distinct spatiotemporal signatures. While tissue moves coherently at cardiac and respiratory frequencies (< 2 Hz), blood flow decorrelates rapidly over time due to the random motion of individual cells. This difference is exploited in eigenvalue-based filtering methods in order to robustly extract the flow signal (*23, 24*).

Extending these principles to volumetric imaging with a miniaturized probe introduces additional constraints. Unlike 2D probes that concentrate acoustic energy within a thin slice using large apertures, our volumetric probe must distribute energy throughout a 3D volume while maintaining SNR and adhering to thermal and mechanical safety limits (*25–27*).

We addressed these constraints through an optimized “wide beam” transmission strategy, similar to (*28*). Rather than using fully unfocused transmissions that disperse acoustic energy throughout the volume, we transmit sixteen sequential beams at 7.2 kHz pulse repetition frequency, each concentrating energy into a 15^◦^ × 15^◦^ sub-volume. Coherent combination of these acquisitions yields volumetric images at 450 Hz across the complete 60^◦^ × 60^◦^ field of view, achieving imaging depths up to 8 cm (Figure 2A-B). This approach achieves sufficient sensitivity while maintaining safety compliance (Table S1).

**Figure 2:**
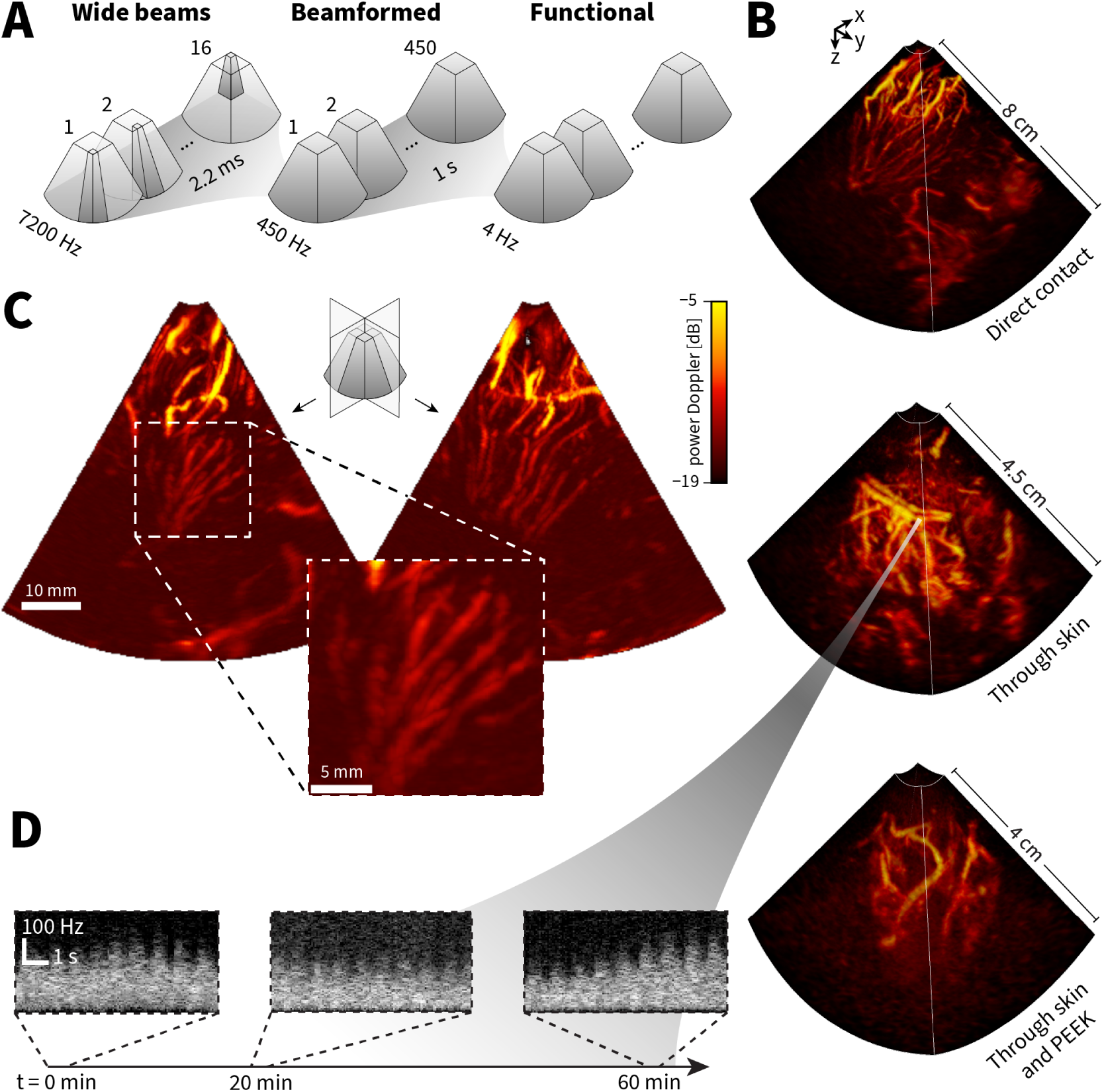
Volumetric hemodynamic imaging performance across three subjects. (**A**) Imaging sequence using 16 wide-beam transmissions at 7200 Hz, reconstructed to 450 Hz volume rate, then filtered to isolate blood flow at 4 Hz for functional analysis. (**B**) 3D renders of power Doppler volumes showing cerebral vasculature in three subjects: direct brain contact during neurosurgery (top, 8 cm depth), through skin-covered cranial opening (middle, 4.5 cm depth), and through skin-covered polyetheretherketone (PEEK) skull prosthesis (bottom, 4 cm depth). (**C**) Orthogonal slices through volumetric data from direct brain imaging, with magnified view showing vessels as small as 800𝜇m diameter. (**D**) Signal stability over one hour showing Doppler spectra from the same 1 mm^3^ region at 0, 20, and 60 minutes.

Real-time hemodynamic feedback is paramount for neurosurgical guidance and emerging closed-loop applications such as brain-machine interfaces (*16, 29*). To meet these demanding requirements, we integrated our probe with a Verasonics research scanner that digitizes the 192 micro-beamformed channels at a rate of 4.2 MHz with 14-bit resolution, generating 1-2 GB/s data throughput. While this probe architecture has been recently tested for general ultrasound applications and 2D ultrafast imaging (*30*), functional volumetric neuroimaging represents a novel application domain requiring specialized 4D processing pipelines for hemodynamic sensitivity and real-time brain activity mapping.

We developed an open-source GPU-accelerated software pipeline to perform real-time volumetric beamforming, eigenvalue-based clutter filtering, and power computation over temporal ensembles of volumes, processing the 450 Hz acquisitions into hemodynamic images at 1-10 Hz for functional analysis (the exact frame rate depends on the chosen ensemble size and overlap). Critically, this pipeline also handles live volumetric rendering to visualize the 3D cerebral vasculature during experiments.

We characterized imaging performance across three experimental conditions relevant to both clinical and research applications: direct cortical contact, transcutaneous imaging through cranial defects, and imaging through a sonolucent skull prosthesis (polyetheretherketone, PEEK), as shown in the bottom image in Figure 2B. Imaging with the probe directly on the brain achieved the highest resolvable depth of up to 8 cm, while transcutaneous imaging achieved depths of up to 4.5 cm (no PEEK) and 4 cm (with PEEK), sufficient for comprehensive cortical mapping. In all cases, vessels with diameters as small as 800 𝜇m are clearly resolved (Figure 2B-C). Furthermore, signal quality remained stable across extended imaging sessions, illustrated by the consistent Doppler spectra measured over one hour (Figure 2D), enabling the lengthy experimental protocols required for comprehensive functional mapping.

The system’s sub-millimeter resolution, large imaging depth and real-time visualization within a miniaturized form factor establish the foundation for functional brain mapping applications demonstrated in subsequent sections.

## Functional ultrasound imaging (fUSi)

The critical question is whether the system’s hemodynamic imaging performance translates into meaningful maps of brain activity. Functional validation requires demonstrating not only that brain activity can be detected, but that the FOV and spatial resolution can resolve the fine-scale organization of cortical networks. To this end, we designed experiments targeting the sensorimotor system, where prior research has established clear relationships between neural activity patterns and hemodynamic responses (*31*) and the well-characterized somatotopic organization of motor and sensory cortices provides an ideal testbed for evaluating spatial precision (*32–36*). Furthermore, the three-dimensional structure of sensorimotor networks, exhibiting complex spatial relationships, specifically requires the volumetric imaging capabilities that distinguish our approach from conventional 2D fUSi.

In our first subject, who had undergone cranioplasty with a sonolucent PEEK implant following hemicraniectomy, we mapped sensorimotor responses during a lip licking task (Figure 3). Posttrauma, this subject suffered from severe aphasia but only minor motor deficits. Using a custom helmet for probe positioning as in our previous work (*13*), we recorded functional responses through the subject’s skin and underlying PEEK prosthesis. Despite the acoustic interface losses, we successfully captured orofacial activation patterns that corresponded with our previous 2D fUSi findings, fMRI data from a closely related orofacial task we previously acquired in this subject (*13*), and the expected location for the lips in the somatotopic homunculus (*37, 38*). The 4D fUSi data revealed activation patterns with high spatial and temporal detail, demonstrating that hemodynamic responses can be resolved even in response to short stimuli (Figure 3C). In line with previous work (*12, 13*), these findings demonstrate the feasibility of fUSi through acoustically favorable implants.

**Figure 3:**
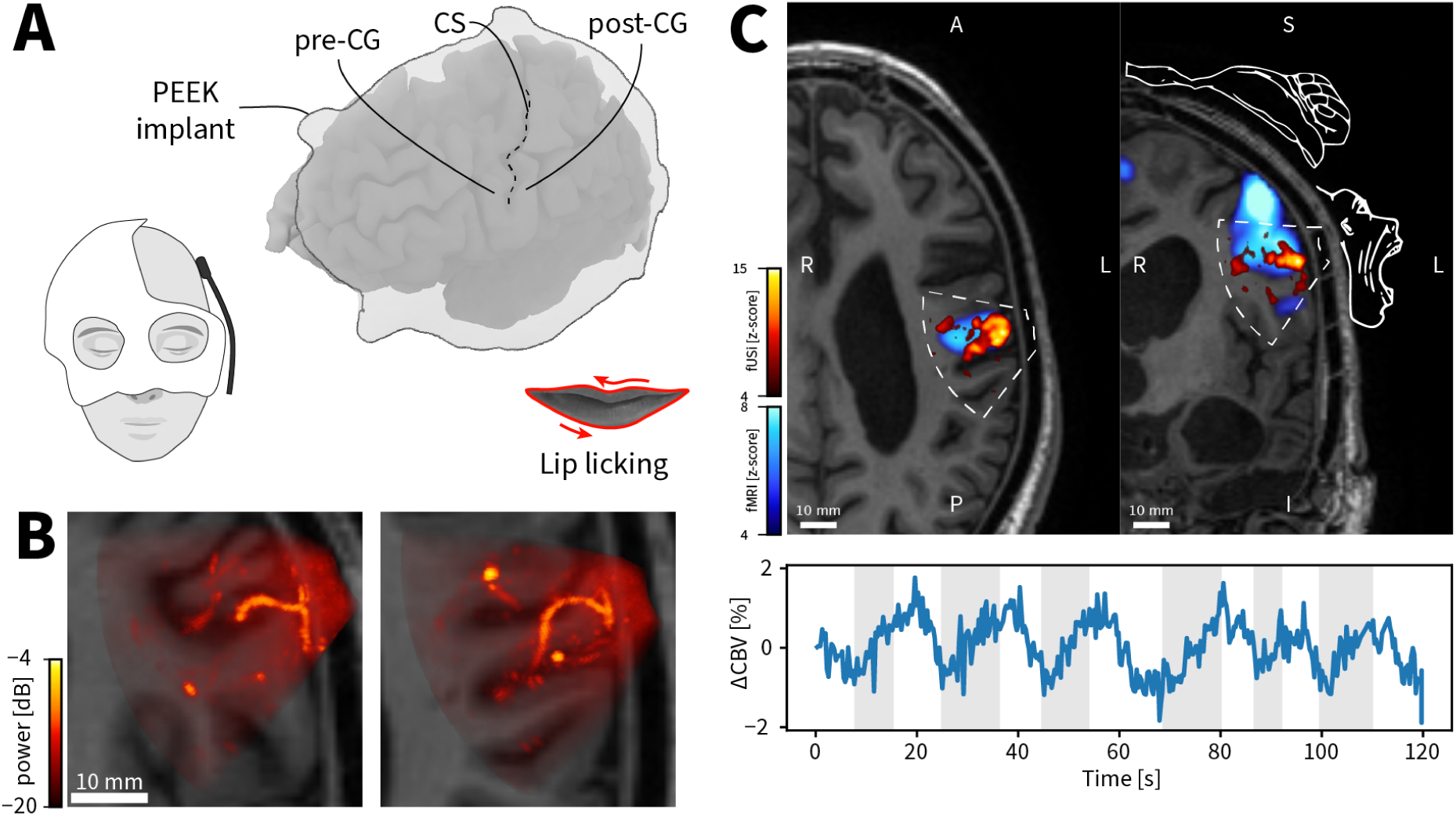
Functional brain mapping through a sonolucent skull prosthesis. (**A**) Subject wearing custom helmet-mounted probe (left) and 3D rendering showing PEEK implant location and imaged brain regions including precentral (pre-CG) and postcentral (post-CG) gyri around central sulcus (CS) (right). (**B**) Co-registered power Doppler volumes (orthogonal 5 mm maximum projections) overlaid on subject’s anatomical MRI. (**C**) Somatosensory activation during lip-licking task. Top: Activation maps from 4D fUSi (red, 𝑝 < 0.01) and prior 3D fMRI (blue, 𝑡 > 4) overlaid on anatomical slices, with homunculus schematic showing expected lip location. Bottom: Average cerebral blood volume (CBV) time course from significantly activated voxels, showing ∼ 2 % signal change during task blocks (gray bars).

Our second subject had a cranial defect overlying the right central sulcus, resulting from the removal of an infected bone flap following craniotomy for a small meningioma. This subject was neurologically intact with no postoperative deficits, enabling more complex functional mapping experiments to evaluate the system’s capability for detailed localization.

Using a custom helmet for stable probe positioning co-registered to the subject’s MRI, we performed systematic motor mapping experiments while the subject moved individual fingers of their left hand or toes of either foot (Figure 4A). Volumetric activation maps computed using a conventional general linear model (GLM) revealed the expected somatotopic organization: foot movements activated medial cortical regions while finger movements engaged the lateral “hand knob” of the precentral gyrus, with distinct patterns for motor output in precentral and sensory feedback in postcentral regions (Figure 4B) (*37, 39*).

**Figure 4:**
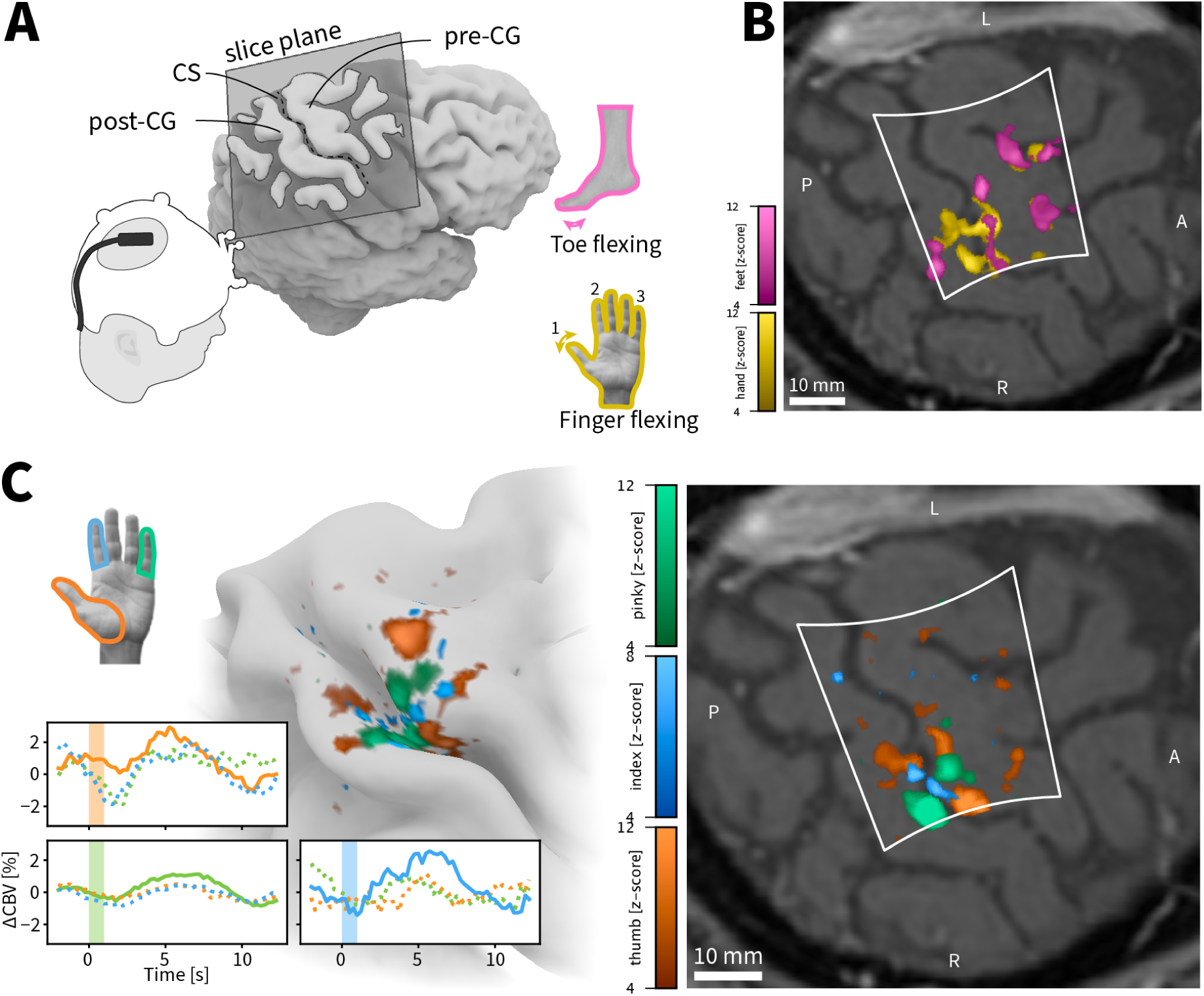
High-resolution somatotopic mapping of the sensorimotor cortex. (**A**) Subject with cranial defect over right motor cortex wearing probe-mounted helmet (left) and oblique slice orientation used for visualization (right). (**B**) Differential somatotopic organization from finger flexion (yellow) and toe flexion (magenta) tasks, showing expected medial foot and lateral hand representations (all contrasts 𝑝 < 0.01, cluster > 100 voxels). (**C**) Fine-scale digit mapping within hand area. Selective contrasts identify voxels preferentially responding to thumb (blue), index (orange), or pinky (green) movements (all contrasts 𝑝 < 0.01, cluster > 100 voxels). Left: Surface projection smoothed to visualize central sulcus. Right: Oblique slice view. Bottom: Average CBV time courses from voxels with highest selectivity for each finger, aligned to movement onset.

The spatial resolution of 4D fUSi enabled clear differentiation between representations of individual digits. Selective contrasts for thumb, index, and pinky movements revealed distinct but partially overlapping activation clusters spanning both the motor and sensory cortex (Figure 4C). Each finger produced a unique spatial signature within the hand area, with activation patterns consistent with high-resolution fMRI studies (*32–36, 39*). This fine-scale mapping demonstrates that 4D fUSi captures the three-dimensional structure of sensorimotor networks, revealing spatial relationships difficult to reconstruct from 2D imaging planes.

To test whether the system’s spatiotemporal resolution could enable real-time applications, we challenged our system with single-trial decoding of naturalistic movements. The same subject played simplified piano melodies (left hand only) on a MIDI keyboard while wearing the probe-helmet. Our aim was to predict, for each of the five fingers whether it moved at any given time, using only the subsequent seven-second window of volumetric hemodynamic data without trial averaging. We trained a 4D spatiotemporal Vision Transformer, commonly used in video recognition problems (*40–42*), leveraging fUSi’s high spatial resolution and sampling rate to learn relevant features from the 4D image data (Figure 5A). After training on four datasets comprising about 15 minutes of acquisitions, the model accurately detected individual finger presses in an unseen melody, confirming that our 4D volumes contain sufficient fidelity for single-trial decoding. Figure 5B illustrates that the model’s learned attention map (*43*) focuses on physiologically relevant regions (primarily the hand-related somatosensory cortex) and peaks at ∼ 3 − 4 seconds after segment onset, which is consistent with the expected hemodynamic delay. Remarkably, these patterns emerged without any anatomical priors or assumptions about the hemodynamic response function, validating both the system’s spatiotemporal imaging quality and the model’s ability to learn neuroscientifically relevant features.

**Figure 5:**
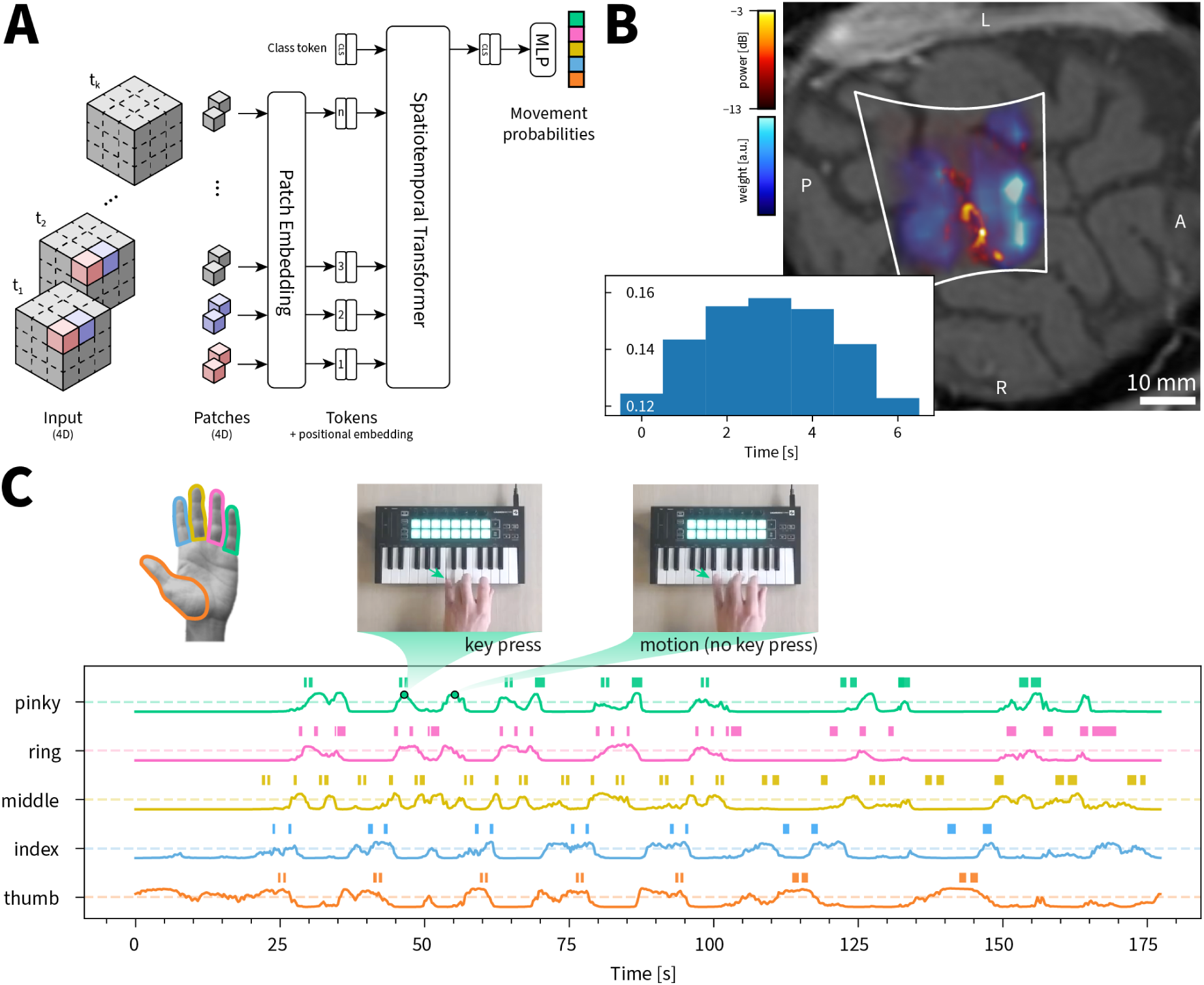
Single-trial decoding of individual finger movements during piano playing. (**A**) Architecture of spatiotemporal Vision Transformer model. Input volumes (28 frames = 7 s at 4 Hz) are divided into 4D patches, encoded with positional information, and processed through transformer blocks to predict finger movement probability for each digit. (**B**) The model’s learned attention maps averaged across test trials, indicating brain regions and time points most informative for movement detection. Top: Spatial attention (blue colormap) overlaid on power Doppler anatomy, highlighting somatosensory cortex including central sulcus (CS). Bottom: Temporal attention profile peaking at ∼ 3 s, consistent with hemodynamic response timing. (**C**) Real-time movement predictions on held-out piano melody. Continuous probability outputs for each finger with 0.5 decision threshold (dashed lines). True key presses shown as colored markers above each trace.

The combination of volumetric coverage, millimeter-scale spatial resolution, and sufficient SNR for single-trial analysis addresses key limitations of current portable neuroimaging methods, particularly for applications requiring real-time feedback or imaging during natural behaviors.

## Discussion

Our development of a miniaturized 4D functional ultrasound imaging system addresses a critical gap in neuroimaging: the need for portable, high-resolution brain monitoring that operates in real-world environments. Achieving sub-millimeter spatial resolution at 1-10 Hz functional frame rates from a device with a footprint of 11.52 × 8.64 mm, the system bridges this gap in a form suitable for diverse clinical and research applications.

The technical foundation of our approach (integrating 3072 elements with a custom tileable ASIC for 16:1 channel reduction) represents a scalable solution to volumetric ultrasound imaging. Miniaturization introduces important engineering constraints: reduced aperture decreases signal-to-noise performance while compact footprint limits heat dissipation, constraining transmittable power. We addressed these challenges through optimized “wide beam” transmission sequences and direct ASIC amplification to maintain adequate sensitivity within safety constraints (Figure 2, Table S1).

This work demonstrates volumetric fUSi feasibility rather than validating functional ultrasound itself, as previous studies have established fUSi’s ability to detect brain activity (*7–12, 14, 44*). Our validation across three clinical subjects (Figure 2) required existing skull openings or acoustically favorable implants, constraining accessibility compared to transcranial techniques. The system achieves 5 cm imaging depth through cranial windows or sonolucent implants (8 cm with direct cortical contact), enabling detailed cortical mapping, but not fMRI’s whole-brain coverage, representing a fundamental trade-off between portability and field of view. Temporally, the physiological ∼ 3 − 4-second hemodynamic response delay imposes constraints on rapid feedback applications (*12, 17*).

Nevertheless, our results establish that 4D fUSi captures physiologically meaningful brain activity with superior spatial detail compared to current portable methods. Single-trial decoding of finger movements during naturalistic piano playing (Figure 5) demonstrates sufficient signal quality for brain-computer interfaces, although the hemodynamic delay necessitates careful system design. The decoding model learned attention patterns focused on somatosensory cortex without anatomical priors, validating imaging fidelity and biological plausibility.

Volumetric hemodynamic imaging enables investigation of three-dimensional activation patterns that sequential 2D scanning cannot capture, particularly important for sensorimotor integration spanning multiple cortical areas. Our somatotopic mapping (Figure 4) revealed distinct but overlapping digit representations consistent with high-resolution fMRI studies (*35, 39*).

Clinically, this technology could significantly impact neurosurgery through continuous hemodynamic monitoring, enabling safer tumor resection and more precise aneurysm or arteriovenous malformation treatment. Real-time feedback helps surgeons make informed vessel intervention decisions, potentially reducing post-surgical deficits (*45*). Applications extend to traumatic brain injury monitoring and neonatal care through the fontanel (*14*). While rigorous comparison with established functional mapping methods—direct electrical stimulation, intra-operative neurophysiology, and high-resolution fMRI—are required for solid clinical validation, our results demonstrate feasibility across relevant scenarios.

The technology’s portability creates new research opportunities for studying the brain in natural environments. Brain-machine interface applications benefit from volumetric coverage and real-time feedback, as demonstrated by successful finger movement decoding, though hemodynamic timing considerations are essential for closed-loop applications. The probe’s small size enables potential subcranial implantation between skull and dura with PDMS encapsulation (*46*), providing chronic hemodynamic access where repeated monitoring is valuable.

Our work demonstrates that miniaturized 4D functional ultrasound imaging represents a significant technological advancement with clear practical applications. The convergence of ASIC technology, optimized sequences, and real-time processing creates unprecedented portable brain monitoring opportunities. While accessibility and validation challenges remain, the demonstrated capabilities establish 4D fUSi as a valuable neuroimaging technology that overcomes longstanding constraints on observing the brain’s intricate dynamics across space and time.

## Data Availability

All data produced in the present study are available upon reasonable request to the authors.

## Acknowledgments

We thank the Neurosurgery OR team and the radiology team at Erasmus MC for facilitating measurements during surgery and fMRI acquisition, respectively. We are grateful to the three study participants for their time and participation.

## Author contributions

L.V., S.S., and P.K. formulated the study design. A.J.F., B.L., A.J.J., N.J.R., and M.B. designed and built the probe hardware. L.V., A.J.F., B.L., A.J.J., N.J.R., and M.B. integrated the probe with the imaging system and optimized probe configurations for human imaging. L.V. developed software for real-time imaging. L.V., J.V., and P.K. developed imaging sequences. S.S. and A.J.P.E.V. handled subject recruitment and clinical oversight. A.J.P.E.V. conducted intraoperative scanning. S.S. and G.S. designed the probe helmets. S.S. acquired fMRI data. L.V., S.S., and P.K. acquired fUSi data. L.V., J.V., and P.K. performed general data analysis. L.V., S.S., J.V., and P.K. interpreted the results. L.V., S.S., J.V., and P.K. wrote the initial manuscript. All authors reviewed and approved the final manuscript.

## Data and materials availability

All data and relevant source code will be made available upon publication.

## Materials and Methods

### Probe Design and Fabrication

The presented ultrasound device, manufactured by Oldelft Ultrasound (Delft, Netherlands), consists of a matrix of 64 × 48 (3072) piezoelectric transducer elements, with a 180 𝜇𝑚 × 180 𝜇𝑚 pitch, connected to three application-specific interface circuit (ASIC) tiles. The transducers have a -6 dB bandwidth of 60% with a center frequency of 4.72 MHz. The resulting composite waveform can be seen in Supplemental Figure S3.

Each ASIC tile can interface with 64 × 16 transducer elements (architecture visualized in Supplemental Figure S1). Each element has its own pulser, time gain compensation (TGC) and low noise amplifier (LNA) circuit. Sets of 4 × 4 elements are connected in a group which performs the receive micro-beamforming. The output of each micro-beamformer is fed to a dedicated cable driver which disconnects the load of the 48 AWG micro-coax cable from the micro-beamformer circuitry. Subsequent subsections will go further in detail on the transmit, receive, and control capabilities of the ultrasound device.

The ultrasound device used in the study consists of three ASICs placed next to each other along each ASIC’s long axis to form an array of 3072 (64 x 48) elements and 192 output groups, and thus only requiring 192 channels from the ultrasound console. The total aperture equals 11.52 mm x 8.64 mm.

#### Transmit

The integrated transmit circuitry contains an on chip pulser capable of transmitting voltages up to 100 V. It also contains a waveform generator, is capable of binary apodization, and has element-level control of delays.

Waveform generation is accomplished using a per-element two level pulser and a versatile waveform generator. The waveform generator has a waveform memory of 16 bits and allows for repeats of parts of the waveform memory to achieve longer pulses. The time quantization of a bit in the waveform 𝑡_s_ can be programmed between 20 and 50 ns with a step size of 1.25 ns. Allowing for transmit frequencies for the full bandwidth of the ultrasound device.

The delay range of each individual element in the transmit beamformer depends on the same transmit clock divider and can span minimally from 0 to 290 ns, to maximally from 0 to 725 ns. The delays are split up in 30 steps where one unit is, 𝑡_s_/2 and step 0 is no delay. On top of the individual element delays a delay can be applied on a group level, with 255 steps with the full step size of the transmit waveform, giving a max range of 255 × 50 × 10^−9^ = 12.75 𝜇𝑠.

#### Receive

In receive mode, sets of 4×4 elements are connected in a “micro-beamforming” group, where a programmable fixed or dynamic focus delay-and-sum beamforming step is performed on the ASIC, reducing the 16 element signals down to a single output channel. The delay range of each individual element in the receive micro-beamformer spans from 60 to 480 ns over 23 steps with quantization steps of 20 ns.

TGC can be applied at the element level with a range of 0 to -20 dB in steps of 2.5 dB. The timing of TGC steps is applied, in the time domain, on a range of 1.28 to 9.60 𝜇s in steps of 640 ns with a possible extra offset with a range of 0 to 19.2 𝜇s and steps of 1.28 𝜇s.

#### Control

A dedicated field programmable gate array (FPGA) (XC7A100T (AMD, Santa Clara, USA)) controls the ASIC tiles during sequencing, uploading transmit and receive micro-beamforming delays, and providing timing control using an on-board 100 MHz clock that is phased locked with the reference clock of the ultrasound console for phase accurate timing and synchronization of the hardware event loops.

When cycling through transmit/receive events in an imaging sequence, delays need to be written to the ASIC registers for each element, which can be prohibitively slow for high volume rate imaging. For this purpose, a polynomial compression scheme is used, where decompression on the ASICs allows for efficient writing of element-wise transmit/receive delays. Third-order and second-order polynomials are used in transmit and receive, taking 6.8 𝜇s and 6.2 𝜇s for decompression (of which 0.7 𝜇s is for communication), respectively. Thus, allowing for high volume rate acquisition sequences.

#### Probe Fabrication

A high density interconnect (HDI) printed circuit board (PCB) connects the ASIC to an interposer. The PZT (CTS 3265HD) is prepared with a conductive matching layer which is cast directly on the piezoceramic. This matching layer consists of a silver filled epoxy to create an electrical connection between the elements and the ground foil. The PZT with matching layer is then applied to the ASIC assembly, on top of the interposer, using the same silver filled epoxy. The elements are diced with a 30 𝜇m dicing blade using a dicing machine (DISCO DAD3241, Tokyo, Japan). Finally, an aluminium foil layer, which acts as the ground electrode for the full array, is applied on the array of elements and connected to a ground pad of the PCB.

This full assembly is then assembled on an aluminum heat sink, to evenly spread the heat generated during imaging over the complete surface area of the tip. The connection between ASIC assembly and heatsink is made using a proprietary epoxy mixture that dampens the sound waves going to the backside as well as creating a thermal conductive layer to transfer the heat from the ASIC to the heatsink. A thermistor is placed within the heatsink to monitor tip temperature during imaging. The acoustic module with heat sink is assembled in its PPSU housing and the acoustic window is cast directly on the array using RTV-J 6130 (Silastic). A microscopy photo of the assembled probe can be found in Figure S2.

### Acoustic Characterization and Safety

Characterization of the acoustic field was performed by submerging the probe’s acoustic tip in a water tank and positioning a calibrated hydrophone (200 µm, Precision Acoustics Ltd, United Kingdom) on a computer-controlled stage (Zaber Technologies Inc., Canada) to sample 128 by 128 points in the 𝑥𝑦-plane immediately in front of the transducer. The hydrophone was driven laterally in 200 µm steps along x and y while triggering a 15° diverging pulse; the received waveform was digitized at 125 MHz for 4096 samples per transmit using a digitizer card (M4i.4421-x8, Spectrum Instrumentation GmbH, Germany). Using the angular spectrum method (*47*) and assuming a tissue propagation model (attenuation coefficient 𝛼 = 0.3 dB MHz^-1^ cm^-1^), the pressure field was propagated to depths up to 80 mm, and from the resulting fields at the global peak location we calculated the mechanical index, soft tissue thermal index, and peak intensity values according to (*25–27*). Thermal testing comprised two 30 min runs: first, a 15° diverging beam was transmitted into a phantom (with pulse repetition frequency of 7.2 kHz) while a thermocouple (USB-TC01, National Instruments, United States) on the probe face recorded surface temperature every second; second, the transducer emitted continuously into still air and self-heating was determined with infrared images taken by a thermal camera (C3, FLIR, United States). The results for these measurements are displayed in Figures S3, S4, and Table S1.

### 4D Ultrasound Imaging Sequence

The transmit and receive beamforming tasks associated with volumetric ultrasound imaging are split between the FPGA/ASICs in the probe, and the ultrasound console system (Verasonics). Transmit beam-formation is handled by the probe’s own FPGA and ASICs, where element-wise delay profiles and pulse-excitation waveforms are pre-programmed onto the probes FPGA memory during initialization. Receive beam-formation is achieved in two stages: 1) using fixed-focus delay-and-sum beamforming for each element in a micro-beamforming group (4x4 elements); followed by 2) console-level beamforming using the 192 (16x12) micro-beamforming group signals. Sequence control and sampling of the ultrasound channel data was performed using a research ultrasound machine (Vantage 256, Verasonics, Kirkland, WA, USA).

While the fixed-focus micro-beamforming step is beneficial in terms of channel-count reduction, the downside is that the effective pitch of the micro-beamforming group “elements” is large (720 𝜇𝑚) relative to the wavelength of the transmitted pulse (430 𝜇𝑚). This results in grating lobes being generated in receive when the console steering angle deviates too far from the fixed-focus used in the first stage beamforming step (*28*). Thankfully these grating-lobes can be suppressed by transmitting a narrow enough beam (∼15°×15°) so as not to insonify the grating lobe regions present in the second receive beamforming stage. Thus, in order to reconstruct a full 60°×60° (azimuth × elevation) opening angle, 4×4 ”wide-beams” were transmitted over an angular grid (−22.5° to 22.5° in steps of 15°, in each direction). The virtual focus for each transmit beam was set at a depth of 33 mm behind the probe, using only the central 48x48 elements in transmit (f-number = 3.8). The fixed-focal depth used in transmit was 60 mm (f-number = 7). The pulse repetition frequency (PRF) was set to 7200 Hz, which resulted in a final frame rate of 450 Hz.

### Imaging Characterization

The Bmode image quality of the proposed ultrasound imaging system was assessed using a multi-purpose ultrasound phantom (Model 040GSE, CIRS, Norfolk, VA, USA). The vertical 100 𝜇m wire column was imaged in both an XZ and YZ orientation to calculate the depth dependent point-spread-function (PSF) along both the azimuthal and elevational axes of the probe (S5.A-C). PSF was quantified using the full-width half-maximum of the beamformed envelope signal across the wires.

The stepped anechoic cylinders were imaged to assess contrast (S5.D-E), using contrast ratio (CR), contrast-to-noise ratio (CNR) and generalized contrast-to-noise ratio (gCNR) (*48*) as quantification methods (S5.F). Dark bands at ±15° occur where beams partially overlap and geometric transmit apodization weighting assumption does not match the actual balance in transmit intensity between beams.

### Real-time Volumetric Power Doppler Imaging

In order to balance the bandwidth requirements across the whole imaging pipeline, received channel data was sampled using Verasonics BW50 sampling mode, which approximates IQ sampling with 50% bandwidth. Chunks of channel data were then beamformed using a custom GPU delay-and-sum beamformer implemented in CUDA (Nvidia RTX 5000). After beamforming, but before returning data to CPU memory, a CUDA implementation of singular value decomposition (SVD) based clutter filtering (*23*) was performed to isolate blood signal from the surrounding tissue-clutter signal. The user could input the tissue cut-off rank using slider on the Verasonics graphical user interface. After SVD clutter filtering, the temporal ensemble (128 frames) was summed to obtain a power-Doppler image of size 60^◦^ × 60^◦^ × 50 mm (sampled at 64 × 64 × 128 voxels), which was visualized in real-time using the visualization tool-kit’s (VTK) volumetric GPU ray-caster. The unprocessed channel data (micro-beamformed only) was recorded in parallel to the real-time image reconstruction, where data was written to a fast solid-state disk (T700 4TB PCIe Gen5, Crucial, ID, USA).

### Clinical Study Design

The three subjects were recruited from the Department of Neurosurgery of the Erasmus MC in Rotterdam.

Subject 1 was included for intra-operative imaging during a neurosurgical procedure, to facilitate real-time visualization and monitoring of cortical vasculature and hemodynamics. The images were acquired while the subject was anesthesized, and the probe placed directly on top of the cortical surface (covered by a sterilized casing containing ultrasound gel).

Subject 2 was recruited in a previous study, and had a clinically implanted sonolucent skull prosthesis (polyetheretherketone, PEEK) post-hemicraniectomy for trauma. The exact measurements and thickness of the PEEK-implant can be found in Supplementary Data 1 of our previous paper (*13*). Post-treatment, the subject experienced severe aphasia but relatively minor motor deficits. This subject was included for functional sensorimotor mapping through the skull prosthesis (covered by skin).

Subject 3 had a cranial defect located directly above the right central sulcus and was included for functional experiments to map sensorimotor responses of finger and toe movements directly over the defect (covered only by skin, no prosthesis). The cranial defect remained after the removal of an infected bone flap following craniotomy for a small meningioma. The subject was neurologically intact with no post-operative deficits.

An overview of the clinical characteristics of these three subjects and the conducted scans can be found in Supplementary Table **??**.

### Ethical Approval

Prior to inclusion, written informed consent was obtained from all patients in line with the Dutch national medical-ethical regulations as formulated by The Central Committee on Research Involving Human Subjects (CCMO) in the Netherlands. The Erasmus Medical Center’s Medical Research Ethics Committees (MREC), located in Rotterdam, the Netherlands, reviewed the three research protocols underlying the current study and ethical approval was given on all three occasions:

1. intraoperative functional Ultrasound (fUS)-imaging during awake and anesthetized neurosurgical procedures (fUS Studie), protocol number MEC-2018-037 (Ethical approval was given on 22nd of January 2020)
2. functional Ultrasound (fUS)-imaging in volunteers with a Skull Bone Defect (SBD), protocol number MEC-2019-0689 (Ethical approval was given on 29th of November 2019)
3. functional Ultrasound (fUS) and fMRI-imaging in patients with a Skull Bone Defect (SBD), protocol number MEC-2022-0087 (Ethical approval was given on 24th of May 2022)

All methods in this study were carried out according to the protocols listed above. All subjects gave permission for the use and publication of their images and data.

### Functional Task Design

The functional paradigms were tailored to each subject based on the cortical region accessible through their specific cranial window and their neurological status.

#### Subject 2 (PEEK implant): Orofacial mapping

In line with our previous 2D fUSi experiments (*13*), Subject 2 performed a lip licking task designed to activate orofacial sensorimotor cortex. The paradigm comprised six trials of voluntary lip licking movements, with each trial lasting 7-11 seconds (randomized duration) followed by rest periods of 8-10 seconds. Task timing was cued by visual instructions displayed on screen.

*fMRI protocol:* To validate the 4D fUSi findings, we compared our results for subject 2 with fMRI data previously acquired as part of our prior 2D fUSi study (*13*). The fMRI acquisition employed a block diagram with 30-second ON-OFF periods, following standard clinical protocols at our institution. During the motor portion of this experiment, the subject performed lip pouting movements following visual cues on a screen during the ON blocks.

#### Subject 3 (skull defect): Sensorimotor mapping

Subject 3 performed three distinct motor tasks to comprehensively map sensorimotor cortex organization:

*Finger flexing task:* Each trial consisted of two sequential flexions of a single finger (left thumb, index, or pinky) in randomized order, totaling ten trials per finger (30 trials total). Subjects were instructed to perform deliberate, isolated movements while avoiding co-activation of adjacent fingers.

*Toe flexing task:* Similar to finger movements, subjects performed two sequential flexions of all toes on either the left or right foot, with ten trials per foot (20 trials total).

For both finger and toe tasks, inter-trial intervals were randomized between 8-10 seconds to minimize expectation effects and allow complete hemodynamic recovery between trials.

*Piano playing task:* This paradigm consisted of two phases: (1) a cued finger movement sequence where subjects pressed specific keys in response to visual cues (used for model training), and (2) naturalistic piano playing where subjects performed simple melodies using only their left hand (used for model testing). Each finger press was recorded via MIDI interface for precise temporal alignment with fUSi data.

### Functional Experiment Setup

Subjects were seated in front of a large screen, and task videos were displayed using custom software for managing synchronization to the ultrasound system. Multiple webcams recorded video streams of the subject’s face and finger movements, which were tracked with MediaPipe (Google LLC, Mountain View, CA, USA) for finger-related tasks. In piano-playing tasks, subjects used a Launchkey Mini MK3 MIDI keyboard (Novation, High Wycombe, UK) connected to GarageBand (Apple Inc., Cupertino, CA, USA) for real-time audio output and MIDI recording. An NDI Polaris Vega optical tracking system (NDI, Waterloo, Canada) captured the positions of the ultrasound probe and each subject’s helmet for subsequent volume registration. Synchronization of all recorded data (video, MIDI, ultrasound, tracking) was conducted offline via software-based timestamps.

### Functional Ultrasound Analysis

Offline functional analysis was carried out by clutter filtering the recorded channel data in ensembles of 448 frames (1 second of data) with a 336-frame overlap (75%), producing power Doppler volumes at 4 Hz. Each volume was reconstructed in the spherical domain spanning 60^◦^ × 60^◦^ × 50 mm and discretized at 64×64×128 voxels. Clutter filtering removed the first 112 ranks (25%) of the data, after which each volume was smoothed with a 1 mm Gaussian kernel to enhance the signal-to-noise ratio. A voxel-wise generalized linear model (GLM) was then fit via ordinary least squares, incorporating standard (task vs. baseline) and relative (e.g., between different fingers) contrasts. Task regressors were convolved with a single-gamma hemodynamic response function (HRF) characterized by a peak delay of 5 seconds and a dispersion of 1 second, excluding the post-stimulus undershoot, based on observations that the typical undershoot seen in fMRI was not evident in our fUS data. Additional regressors included a third-order polynomial to account for baseline fluctuations and five high-variance confound regressors, derived as the first five principal components from the top 2% most variable voxels (*49*), to remove system-related artifacts and intensity spikes due to brief movements. Statistical significance was determined via permutation testing with 10,000 shuffles of the design matrix, applying a threshold of 𝑝 < 0.01 and removing clusters smaller than 100 voxels. To determine a voxel’s preference towards one of the fingers in the finger-related tasks, all relative tests (e.g., thumb > index AND thumb > pinky) were required to be significant. Resulting activation maps were overlaid on anatomical or functional MRI scans for visualization, and cortical surface projections were generated with FreeSurfer (*50*), with the default skull-stripping step replaced by HD-BET (*51*) for datasets exhibiting large anatomical defects. All functional visualizations were produced using custom Python scripts built around Matplotlib (*52*).

### fMRI analysis

The fMRI data for subject 2 was processed offline using Statistical Parametric Mapping (SPM8, Functional Imaging Laboratory, UCL, UK) implemented in Matlab. Images were spatially realigned and co-registered to the subject’s T1-weighted anatomical scan using rigid body transformation. Functional images were smoothed with a 6×6×6 mm^3^ Gaussian filter (FWHM). Analysis employed a general linear model with blocks convolved with the hemodynamic response function, corrected for temporal autocorrelation, and high-pass filtered (128 second cutoff). Motion parameters were included as nuisance regressors. Individual t-contrast images comparing experimental versus control conditions were generated and thresholded at approximately 60% of the maximum t-value (𝑡 > 4). The resulting activation maps were projected onto the 3D T1-weighted image and verified by an experienced neuroradiologist to confirm expected sensorimotor activation patterns, with threshold adjustments as necessary.

### Vision Transformer Model for Detecting Finger Motion

We developed a spatiotemporal Vision Transformer (ViT) (*40, 41*) to decode individual finger movements from 4D fUSi data, a novel application of transformer architectures to volumetric brain imaging sequences. Each input consisted of 28 consecutive power Doppler volumes (7 seconds at 4 Hz), with each volume resampled to 64 × 64 × 64 voxels. The 4D data was divided into spatiotemporal patches of size 8 × 8 × 8 voxels × 4 timepoints, yielding 512 patches per segment. These patches were linearly projected to 256-dimensional embeddings and processed through 5 transformer blocks with 4 attention heads, along with a prepended classification token (Figure 5A). To address the limited training data (approximately 15 minutes of piano playing), we first pretrained the model as a masked autoencoder (MAE) (*42*). During pretraining, 75% of patches were randomly masked and the model learned to reconstruct them, forcing it to learn spatiotemporal hemodynamic patterns purely from context. After pretraining, we replaced the decoder with a 5-way classification head to predict which finger moved.

Given the severe class imbalance (periods without finger movement periods outnumbered movements 10:1), we employed balanced sampling during training. The model was optimized using binary cross-entropy loss with a temporal consistency term to encourage smooth predictions. Layer-wise learning rate decay preserved pretrained features while adapting to the classification task.

To verify biological plausibility, we visualized attention patterns using attention rollout (*43*), computed as:

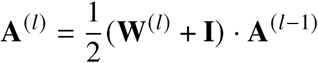

where **W**^(𝑙)^ represents attention weights at layer 𝑙. By marginalizing these 4D attention maps across spatial or temporal dimensions (Figure 5B), we confirmed the model focused on somatosensory cortex with peak attention at ∼3-4 seconds post-movement, matching the expected hemodynamic response. This demonstrates that our 4D transformer learned physiologically meaningful patterns directly from the volumetric data without explicit anatomical constraints. Detailed hyperparameters used during pre-training and fine tuning are provided in Tables S2-S3.

### Volume Registration

To accurately map power Doppler volumes and activation maps to each subject’s anatomical MRI, we employed a passive infrared (IR) optical tracking system (NDI Polaris Vega, Waterloo, Canada; 6 degrees of freedom tracking at 60 Hz). Calibration was essential to determine the spatial transformations between the physical ultrasound probe and the subject’s head. This process involved mounting IR-reflective marker arrays on both the ultrasound probe holder and the subject’s helmet. Fiducial landmarks were first acquired on each object using a tracked stylus, allowing us to capture precise spatial points relative to the reference geometries. These landmarks were then aligned to their corresponding digital models through an iterative closest point (ICP) algorithm, establishing the spatial relationship between the probe and the helmet. Subsequently, the helmet was registered to the subject’s MRI using ICP-based surface matching, with the alignment visually validated in 3D Slicer. Combining the probe-to-helmet and helmet-to-MRI transformations enabled the mapping of ultrasound volumes onto the MRI voxel grid. Minor residual misalignments, typically around 2 mm in translation and 2^◦^ of rotation, were corrected by applying small rigid transforms within 3D Slicer, aligning prominent vascular structures in the power Doppler volumes, such as the central sulcus and pial surfaces, with their anatomical counterparts in the MRI.

---

**Figure S1:**
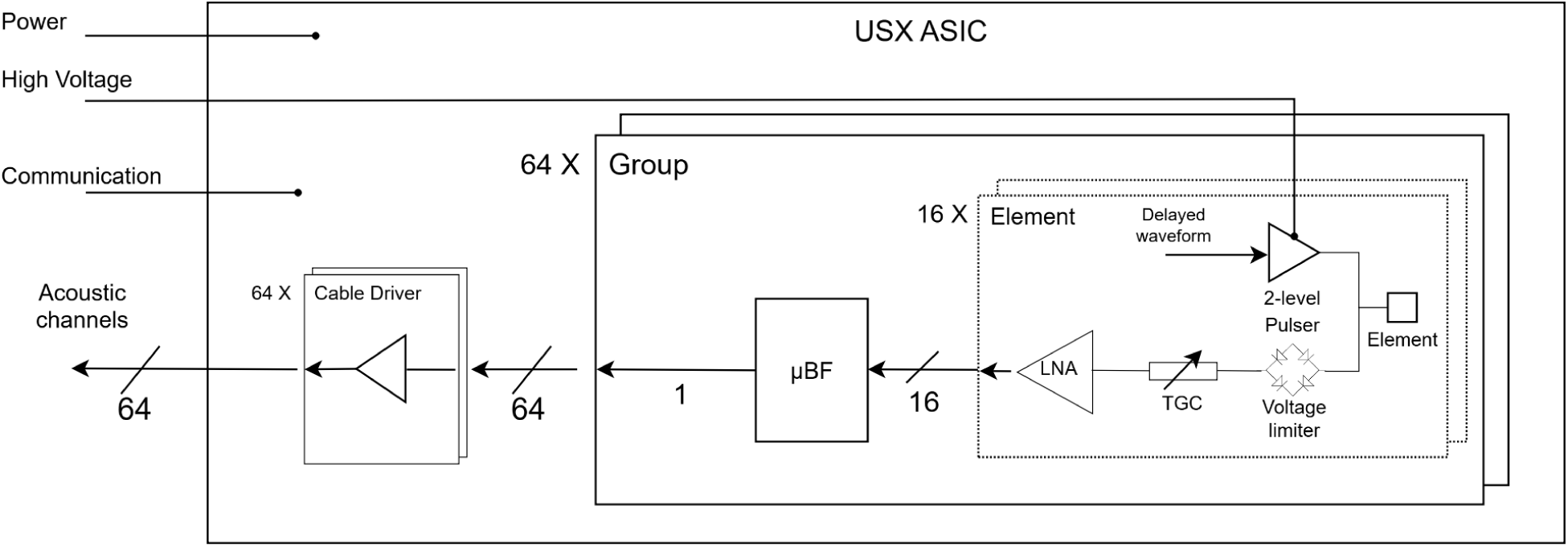
Block diagram of the ASIC circuits.

**Figure S2:**
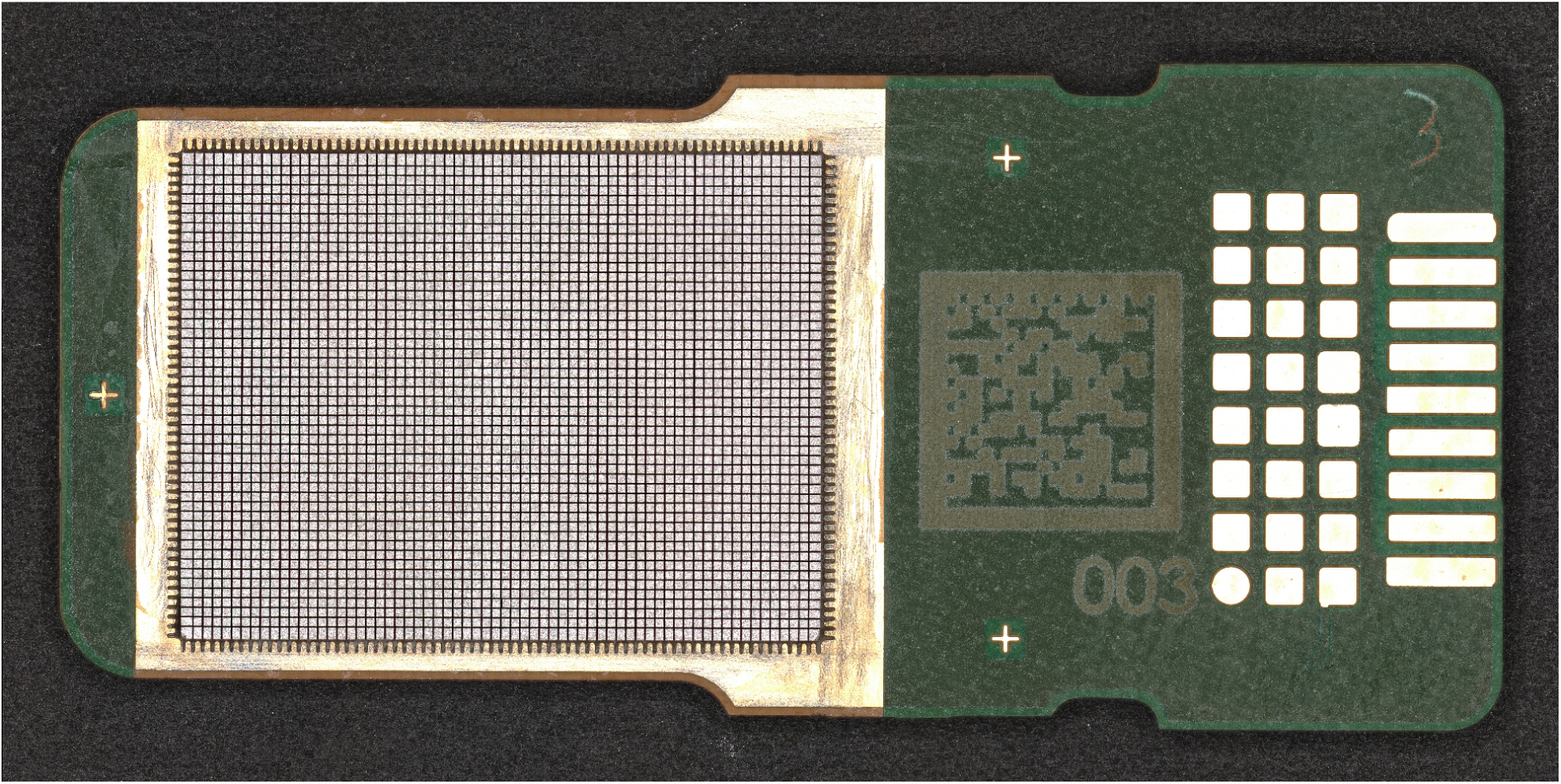
Microscopy photo of the 3072-element probe.

**Figure S3:**
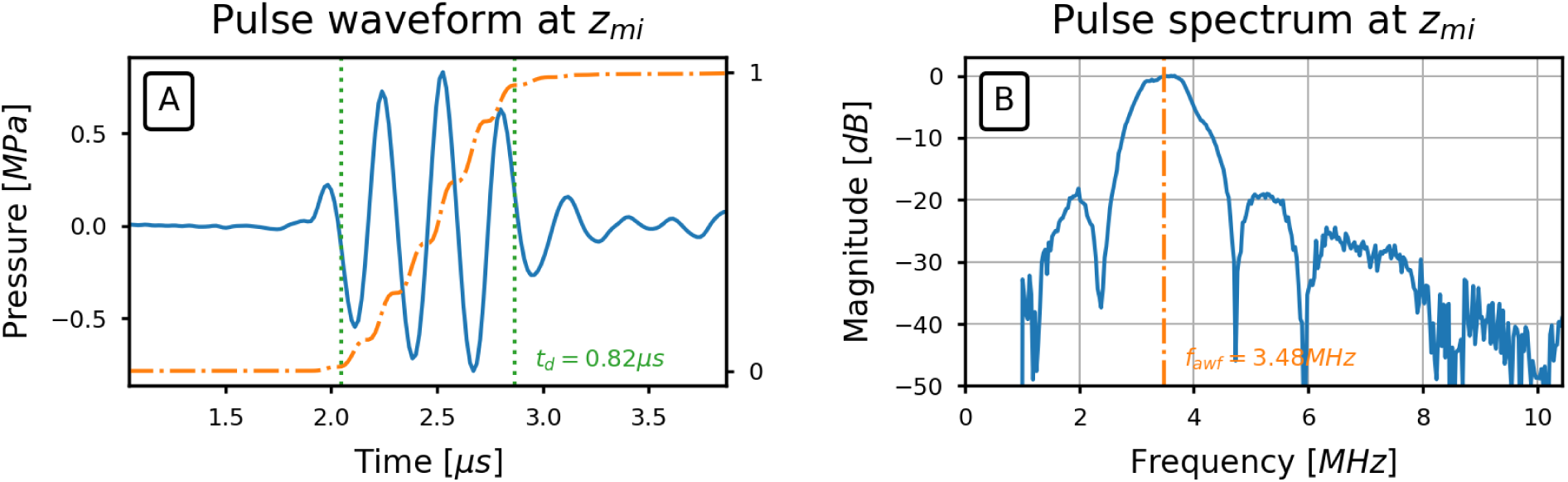
Pressure waveform and spectrum for a 15° wide beam transmission. The measurement was taken at the depth of maximum acoustic intensity, 𝑧_𝑚𝑖_, centered in front of the transducer. The waveform plot also shows the pulse intensity integral, normalized to the final value (orange, dashed line), and the start and end times used for determining the pulse duration (green, dotted lines), defined as 1.25 times the interval between the times when the pressure integral reaches 10% and 90% of its final value. The spectrum plot also shows the acoustic working frequency 𝑓_𝑎𝑤 𝑓_ (orange, dashed line).

**Figure S4:**
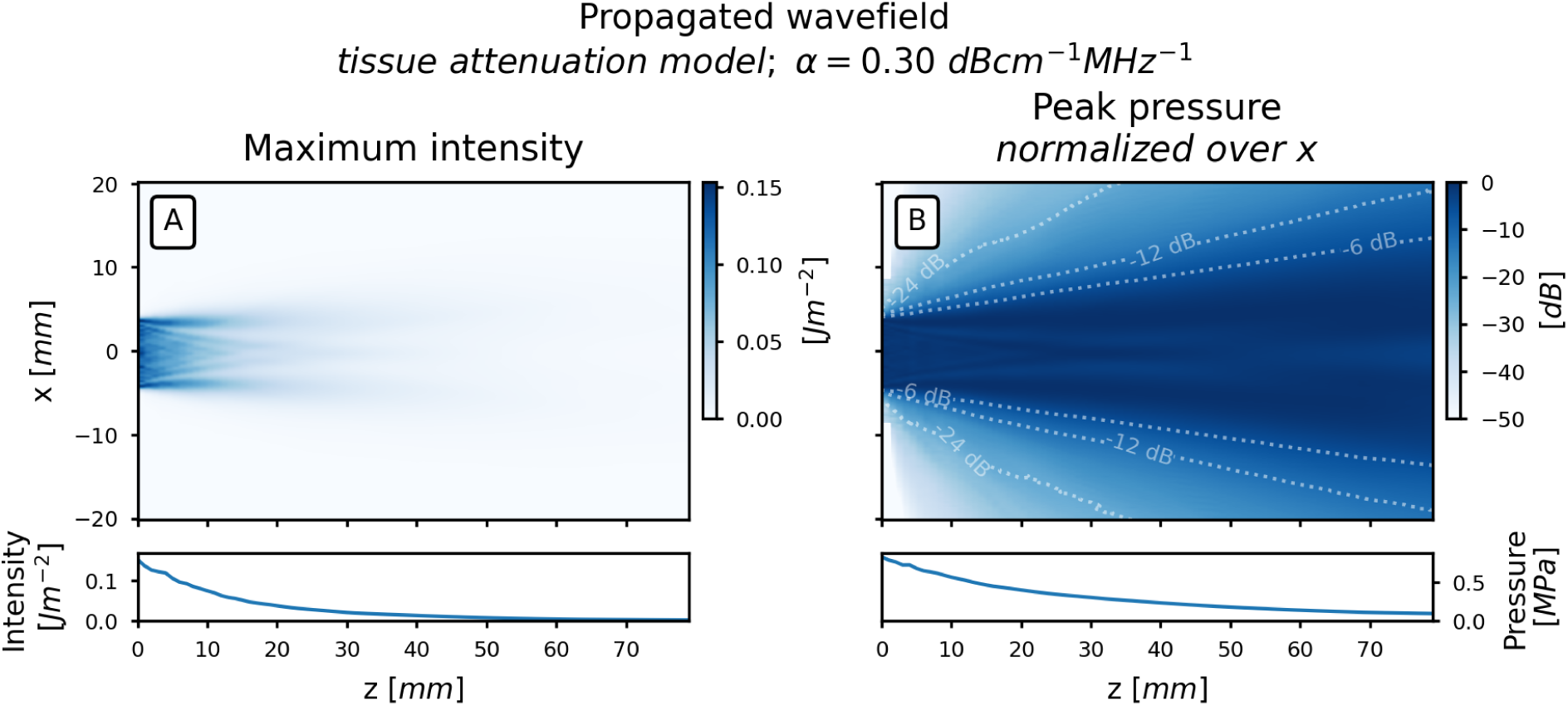
Maximum projection of the acoustic field for a 15° wide beam transmission. The maximum value is shown along the y-axis of the attenuated pulse intensity integral field, 𝑝𝑖𝑖_𝑎_. Each value was calculated by propagating the pressure field in the scan plane at 𝑧 = 0 along the 𝑧-axis with use of the angular spectrum approach (*47*).

**Figure S5:**
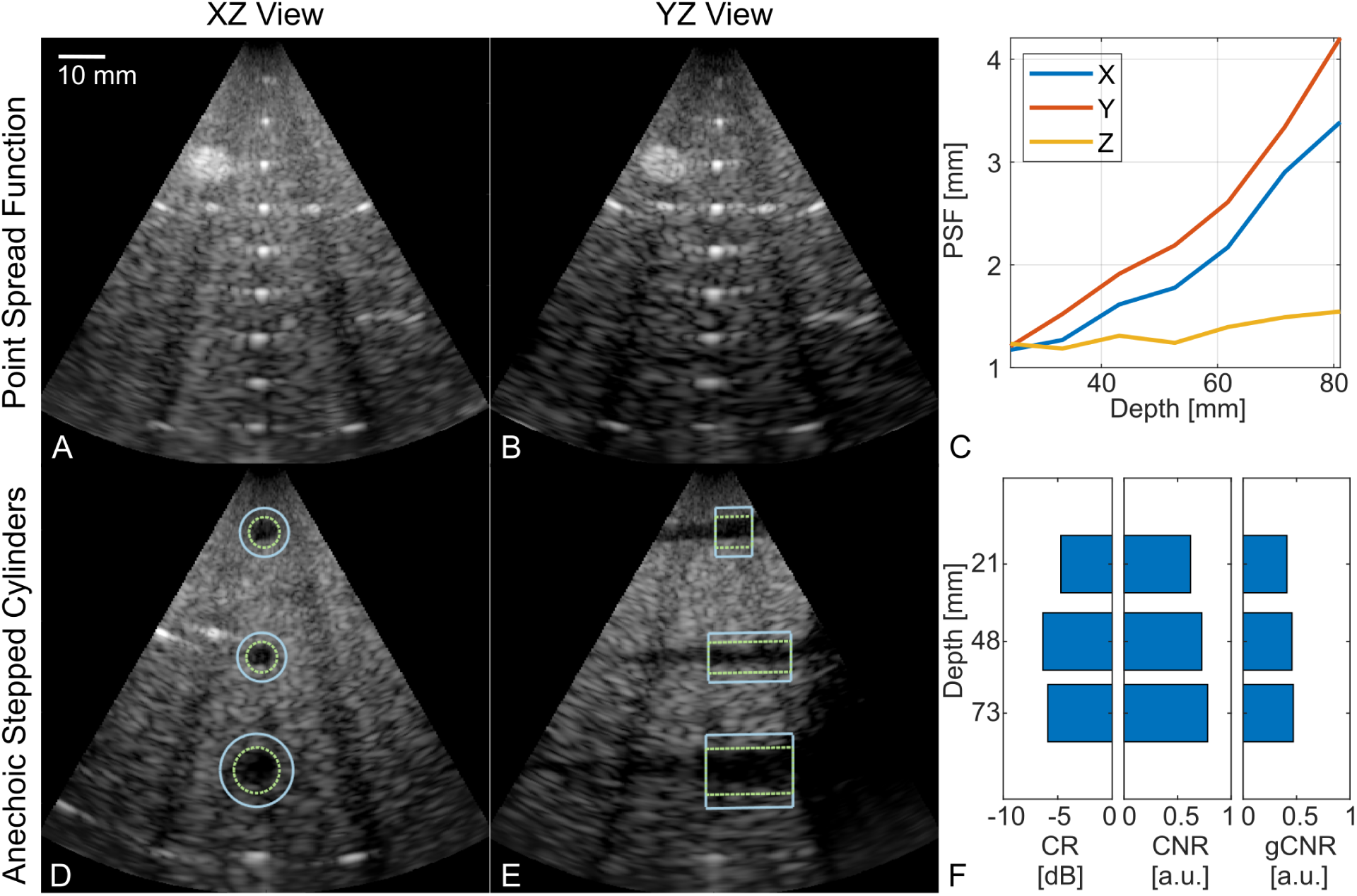
*In vitro* Bmode imaging performance characterization. (A-C) Point-spread-function (PSF) assessment using 100 𝜇m wires vertically spaced 10 mm apart. (D-F) Contrast assessment using anechoic stepped cylinders using contrast-ratio (CR), contrast-to-noise ratio (CNR), and generalized contrast-to-noise ratio (gCNR). (D-E) Green dashed and solid blue paths denote cyst and background regions, respectively.

**Table S1:** Characterization of the Acoustic Transmission Sequence. To verify that the generated pressure field and heating effects fall within established exposure limits, calibration measurements in a water tank were carried out according to (*25–27*).

| Parameter | Units | Measured | Exposure limit |
| --- | --- | --- | --- |
| pulse duration | $\mu\text{s}$ | 0.82 | – |
| working frequency | MHz | 3.48 | – |
| spatial-peak pulse-average intensity, $I_{spp,a}$ | $\text{W}/\text{cm}^2$ | 18.67 | 190 (27) |
| spatial-peak time-average intensity, $I_{spt,a}$ | $\text{mW}/\text{cm}^2$ | 88.20 | 94 (27) |
| mechanical index | – | 0.42 | 1.9 (27) |
| soft tissue thermal index | – | 0.64 | 0.7 (25) |
| max. surface temperature (simulated use, 30 min.) | $^{\circ}\text{C}$ | 30.7 | 43 (26) |
| surface temperature rise (simulated use, 30 min.) | $^{\circ}\text{C}$ | 5.1 | 6 (26) |
| max. surface temperature (still air, 30 min.) | $^{\circ}\text{C}$ | 47.6 | 50 (26) |
| surface temperature rise (still air, 30 min.) | $^{\circ}\text{C}$ | 12.0 | 27 (26) |

**Table S2:** Hyperparameters used during pretraining of the 4D vision transformer model.

| Parameter | Value |
| --- | --- |
| optimizer | AdamW (53) |
| optimizer momentum | $\beta_1 = 0.85, \beta_2 = 0.95$ |
| weight decay | 0.001 |
| learning rate | $5 \times 10^{-4}$ |
| learning rate schedule | cosine decay (54) |
| warmup epochs | 50 |
| epochs | 2000 |
| batch size | 64 |
| gradient clipping | 10.0 |
| augmentation | shift $\sim U(-0.5, 0.5)$ mm, rotation $\sim U(-1, 1)$ degrees |
| masking ratio | 0.75 |

**Table S3:** Hyperparameters used during fine-tuning.

| Parameter | Value |
| --- | --- |
| optimizer | AdamW (53) |
| optimizer momentum | $\beta_1 = 0.9, \beta_2 = 0.999$ |
| weight decay | 0.01 |
| learning rate | $5 \times 10^{-4}$ |
| learning rate schedule | cosine decay (54) |
| warmup epochs | 5 |
| epochs | 100 |
| batch size | 64 |
| gradient clipping | 5.0 |
| augmentation | shift $\sim U(-0.5, 0.5)$ mm, rotation $\sim U(-1, 1)$ degrees |

